# Utilising Artificial Intelligence to Identify Ventricular Tachycardia Ablation Targets in Sinus Rhythm

**DOI:** 10.64898/2026.06.08.26354989

**Authors:** Xuezhe Wang, Joseph Mayer, Adam Dennis, Anthony W.C. Chow, Jaffar Al-Sheikhli, Rafaella Siang, James Winter, Christopher O’Shea, Tarvinder Dhanjal, Pier D Lambiase, Michele Orini

## Abstract

**Background and Aims:** Machine learning has shown potential in predicting ablation targets for ventricular tachycardia (VT) in an animal model. This study progresses to externally validating deep learning approaches for human data.

**Methods:** The development and external validation dataset included 21 and 13 patients, respectively, with structural VT undergoing catheter ablation. In the development datasets, electrophysiological studies were conducted using the Advisor^TM^ HD grid (Ensite^TM^ X), while both CARTO and Ensite Precision were used in the validation dataset.

In each patient, VT ablation targets were defined as mapping points within 8 mm of VT isthmuses. Three advanced machine learning models were trained using cardiac mapping data acquired in both omnipolar and unipolar configurations during sinus rhythm and ventricular pacing. Discrimination was evaluated using nested leave-one-out cross-validation at patient level.

**Results:** Overall, graph convolutional networks (GCNs), which integrate intracardiac signal waveforms with three-dimensional electroanatomical geometries, achieved the highest performance, with optimal results obtained from unipolar electrograms acquired in sinus rhythm (median AUC 0.793, sensitivity 83.6%, specificity 69.0%). This may be partly explained by the inclusion of repolarization dynamics in unipolar electrograms and the higher point density of sinus rhythm maps. Comparable performance was observed in the external dataset.

**Conclusion:** This study demonstrates that graph convolutional networks applied to sinus rhythm EGM waveforms collected during substrate mapping can localise critical components of VT re-entry circuits. This approach has potential to provide fast and accurate ablation guidance without the need to induce and map VT, improving safety and efficacy of VT catheter ablation.

**Structured Graphical Abstract:** Utilising artificial intelligence approaches to identify ventricular tachycardia ablation targets through electrophysiological maps in sinus rhythm.

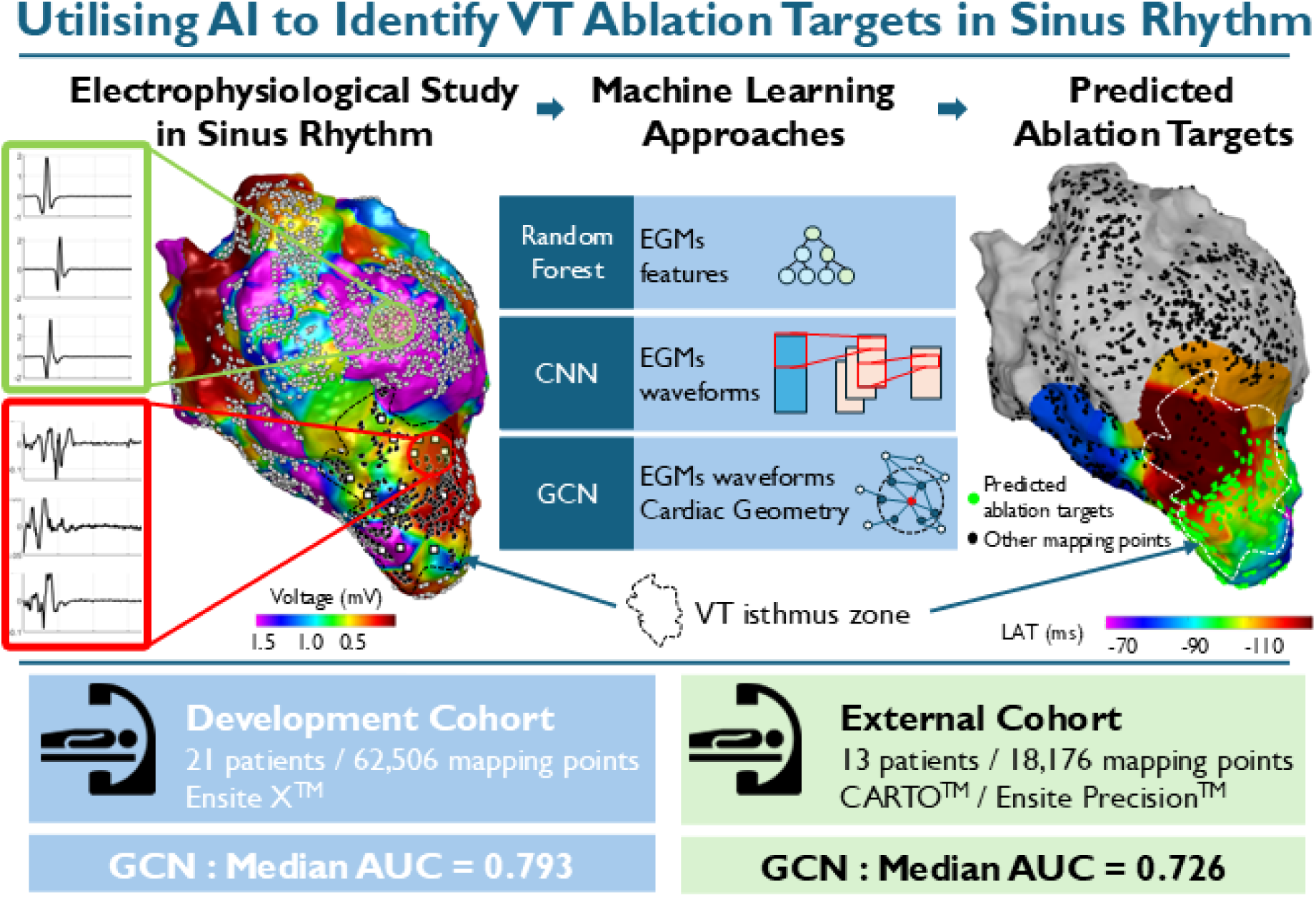

## Introduction

Catheter ablation has become a recommended therapy to control the recurrence of ventricular tachycardia (VT), which aims to terminate the VT through identifying and ablating the critical isthmus of abnormal re-entrant circuits formed around fibrotic tissue ^1,2^. Clinical trials have proven that catheter ablation can significantly reduce the VT recurrence rates compared to medical therapy ^3–5^.

Conventional mapping strategies, such as entrainment and activation mapping, require induction of VT to identify the critical isthmus with VT activation mapping. However, the majority of clinical VTs are haemodynamically unstable ^6^, which limits the feasibility of activation mapping, resulting in unsatisfactory ablation outcomes and increased procedural risk.

To overcome the haemodynamic constraints, substrate-based mapping techniques without VT induction have been proposed and applied ^7^. These approaches can identify the critical VT isthmus through analysing intracardiac electrophysiological data in these substrate maps collected during sinus rhythm or paced rhythm, such as voltage mapping, decrement evoked potential, isochronal late activation mapping and re-entry vulnerability index ^8–12^. However, the pacing protocol and analysis process remain complex and time-consuming, and it is difficult to integrate multi-domain electrophysiological features analysis in real-time ^13^.

Despite recent technological advances, more than half of patients have been shown to experience VT recurrence within one year following catheter ablation ^14^. The rapid development of artificial intelligence technology has demonstrated powerful applications in electrocardiography and in the diagnosis of cardiovascular diseases ^15,16^. Our previous work demonstrated a random forest (RF) approach to predict VT ablation targets with multi-domain features extracted from EGMs in a porcine model ^17^. Despite encouraging performance results, machine learning approaches based on analysis of signal features are limited by the complex feature extraction process, which is heavily dependent on the methodology used to extract features and signal quality ^18,19^.

This study extends the existing RF framework to two retrospective clinical cohorts and proposes more advanced deep learning algorithms that analyse EGM waveforms directly. Advanced deep learning methods, such as convolutional neural networks (CNNs), offer the advantage of learning directly from raw data without relying on predefined features, thereby enabling the extraction of complex signal representations. CNNs have demonstrated considerable potential in cardiology ^20^, particularly when data are organised on a regular grid, such as image pixels or spatially defined leads in the 12-lead ECG. In contrast, graph convolutional networks (GCNs) are specifically designed to capture complex relationships in graph-structured data and may be particularly well suited to the analysis of electroanatomical mapping, where a 3D cardiac map can be represented as a graph of nodes (mapping points) connected by edges encoding anatomical proximity and electrical connectivity.

The aim of this study was to evaluate the potential of graph convolutional networks and convolutional neural networks for automatic identification of ablation targets, and to determine the cardiac mapping configuration (e.g., sinus rhythm versus ventricular pacing, and unipolar versus omnipolar) that maximises model performance. The vision is to deliver a computationally efficient and clinically practical AI-driven solution to identify critical pro-arrhythmic substrate without the need to induce VT.

## Methods

### Electroanatomic mapping

Electrophysiological records of 21 patients who underwent VT ablation at University Hospital Coventry and Warwickshire (Coventry, United Kingdom) between 2021 and 2025 were retrospectively analysed (Table 1). Patients were included in this study only if the VT isthmus was successfully identified with a VT local activation timing map. The study protocol was approved by the local research ethics committee, and all participants gave written informed consent. The ablation procedure was performed with the Ensite^TM^ X system (Abbott Medical, IL, USA) and HD Grid multielectrode catheter (Abbott Medical, IL, USA).

**Table 1.**
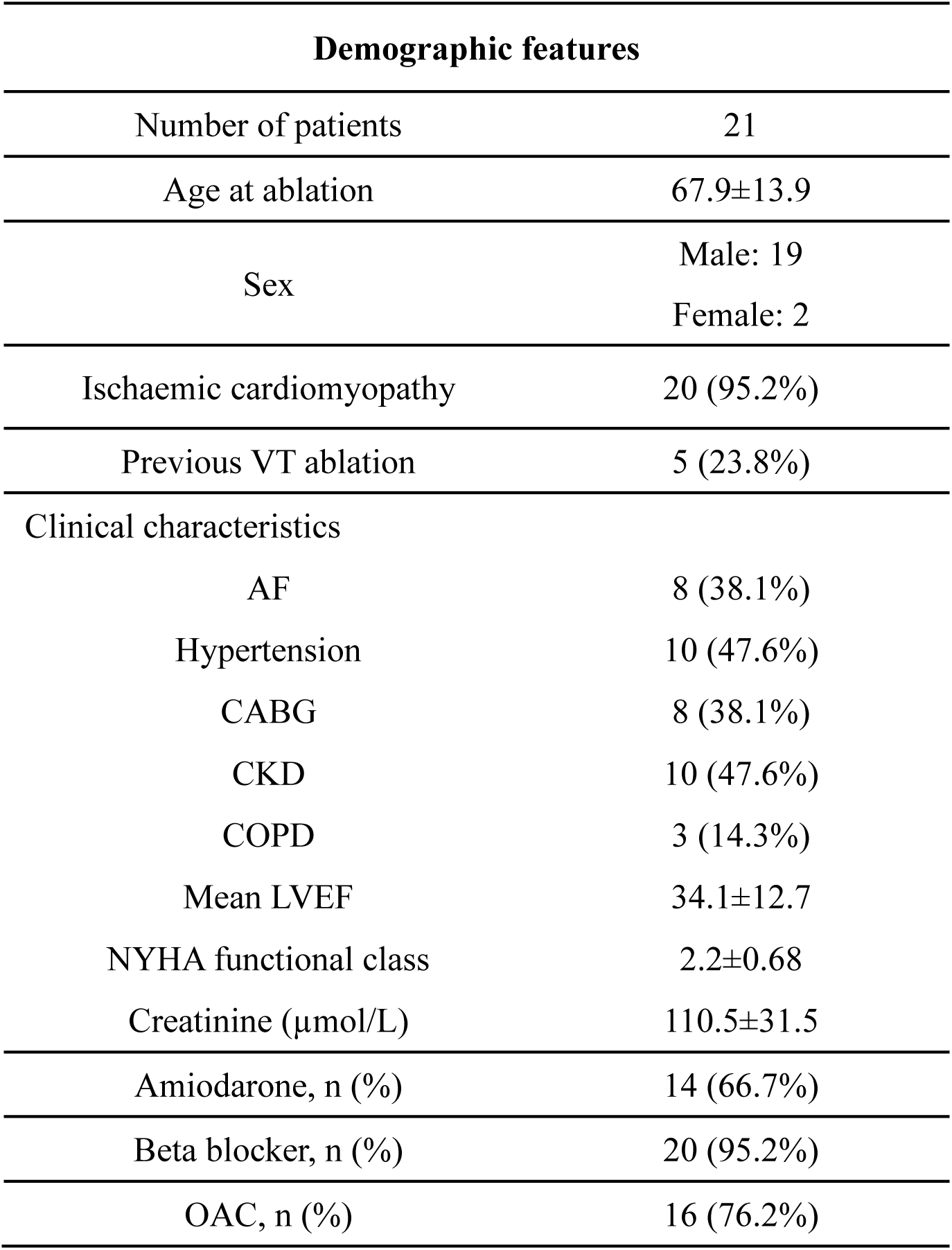
Description of the primary cohort. AF: atrial fibrillation; CKD: chronic kidney disease; COPD: chronic obstructive pulmonary disease; CABG: coronary artery bypass graft; NYHA: New York Heart Association; OAC: oral anticoagulation.

| Demographic features |  |
| --- | --- |
| Number of patients | 21 |
| Age at ablation | 67.9±13.9 |
| Sex | Male: 19<br>Female: 2 |
| Ischaemic cardiomyopathy | 20 (95.2%) |
| Previous VT ablation | 5 (23.8%) |
| Clinical characteristics |  |
| AF | 8 (38.1%) |
| Hypertension | 10 (47.6%) |
| CABG | 8 (38.1%) |
| CKD | 10 (47.6%) |
| COPD | 3 (14.3%) |
| Mean LVEF | 34.1±12.7 |
| NYHA functional class | 2.2±0.68 |
| Creatinine (μmol/L) | 110.5±31.5 |
| Amiodarone, n (%) | 14 (66.7%) |
| Beta blocker, n (%) | 20 (95.2%) |
| OAC, n (%) | 16 (76.2%) |

For each patient, high-density substrate maps were collected within the left ventricle (LV) under two conditions, sinus rhythm (SR), and programmed extra stimulation (ES), during which a drive train at a 600 ms cycle length was delivered from the right-ventricular apex, followed by a single premature stimulus ^12^. The coupling interval of the premature stimulus was set equal to the clinical VT cycle length based on 12-lead ECG or device interrogation ^12^.

Intracardiac EGMs with omnipolar and unipolar configurations were included with a band-pass filter set to 30-300 Hz and 0.5-500 Hz during mapping, respectively. The omnipolar waveforms are generated by combining adjacent unipolar electrodes into local tri-poles, which incorporate bipolar information while offering higher mapping density and reduced sensitivity to wavefront direction ^21,22^. All EGMs were sampled at 2000 Hz. Signal processing tools within the Ensite X platform were applied to suppress noise and attenuate far-field components. Proprietary Ensite X algorithms automatically excluded mapping points with poor signal quality or duplicate waveforms.

The critical VT isthmus was identified from VT local activation time maps, applying the proprietary first deflection omnipolar EGMs annotation algorithm (N=13, 61.9%). Haemodynamically unstable VTs were terminated, and critical VT sites were identified by pace mapping within deceleration zones (N=8, 38.1%). A successful pace map was defined as a >95% 12-lead QRS morphology match to the clinical VT, with latency >40 ms. Multiple VT critical sites were annotated within the mapping system, and an 8 mm radius region around each site was defined as the critical VT isthmus zone. All annotations were verified by expert electrophysiologists. Mapping points located within these annotated VT isthmus zones were labelled as potential VT ablation targets (positive class), the rest of mapping points were considered as non-VT related targets (negative class).

### Signal Processing

Mapping data was reviewed offline using a custom MATLAB graphical user interface (MathWorks, MA, USA). The window of interest was defined for each patient to capture the correct beat (SR and premature extra stimulus, S2, in the Extra Stimulus map) in both omnipolar and unipolar EGMs. The window onset was set immediately after the pacing artefact and before the onset of local activation. For SR maps, the window offset was 500 ms from the onset, while for ES maps, a shorter duration of 400 ms was used to include the S2 beat only. All EGM segments with ectopic signal were excluded. The distribution of mapping points and VT ablation targets for each patient is described in Table 2.

**Table 2.** Distribution of mapping points and VT ablation targets for each data modalities, the data are reported as median (IQR) and the total number N. The VT ablation targets were defined as mapping points located in the critical VT isthmus zone using a radius(R)= 8 mm around the VT critical sites.

|  | **Sinus Rhythm map** |  | **Extra Stimulus map** |  |
| --- | --- | --- | --- | --- |
|  | Omnipolar | Unipolar | Omnipolar | Unipolar |
| Number of maps | 21 | 21 | 21 | 21 |
| Mapping points per map | 3,982<br>(2,594-5,080) | 2,869<br>(2,111-3,879) | 1,368<br>(1,070-1,987) | 608<br>(473-996) |
| Total mapping points | 80,783 | 62,506 | 35,122 | 16,256 |
| VT ablation targets per map (R=8) | 317<br>(161-579) | 251<br>(138-498) | 186<br>(139-299) | 96<br>(64-132) |
| Percentage of VT ablation targets per map (R=8) | 7.90%<br>(5.8%-14.0%) | 8.73%<br>(6.0%-14.4%) | 14.49%<br>(7.1%-22.5%) | 14.43%<br>(7.4%-21.8%) |
| Total VT ablation targets | 7,934 | 6,601 | 5,082 | 2,296 |

Feature extraction was performed using the pipeline previously described ^17^. Band-pass filters at 0.5-20 Hz and 0.5-40 Hz were applied for unipolar and omnipolar EGMs to measure the QRS complex, respectively. Features from functional, spatial, spectral, and time-frequency domains were extracted (20 for omnipolar and 24 for unipolar EGMs, with the latter including 4 repolarisation features, which are not available from the analysis of omnipolar EGMs). Functional domain included local activation and repolarisation time (LAT and RT), activation recovery interval (ARI, a surrogate for local action potential duration ^23^), EGM duration and amplitude, number of deflections and signal changing rate. Spatial domain included spatial gradients of LAT, RT, and ARI. The time-frequency domain included the signal energy from 4 time-frequency bands in each signal channel. The spectral domain included the central frequency and the number of spectral peaks. Detailed feature definitions and computation are provided in the supplementary Table 1.

Both omnipolar and unipolar EGMs were divided into two segments (supplementary Figure 1), the ventricular activation period (segments within the QRS complex) and the post-QRS interval. Functional and time-frequency domain features were calculated separately in both segments to capture distinct features of conduction and repolarisation.

### Model development

The analytical workflow is shown in Figure 1. Three models were implemented: a feature-based RF model, which was used as a benchmark, a one-dimensional convolutional neural network (1-D CNN) and a graph-convolutional network (GCN) ^24,25^. The raw EGM waveforms were used as the input for the neural networks, while signal features constituted the input to the RF model. The RF classifier was achieved using the *TreeBagger* function in MATLAB 2022b. Deep learning models were implemented in Python 3.10 with PyTorch 2.6.0 and PyTorch Geometric 2.6.1. The detailed model architecture, parameters, and hyperparameter ranges are described in supplementary materials. This study was reported with reference to the TRIPOD+AI guidance for clinical prediction models using artificial intelligence or machine learning methods^26^.

**Figure 1.**
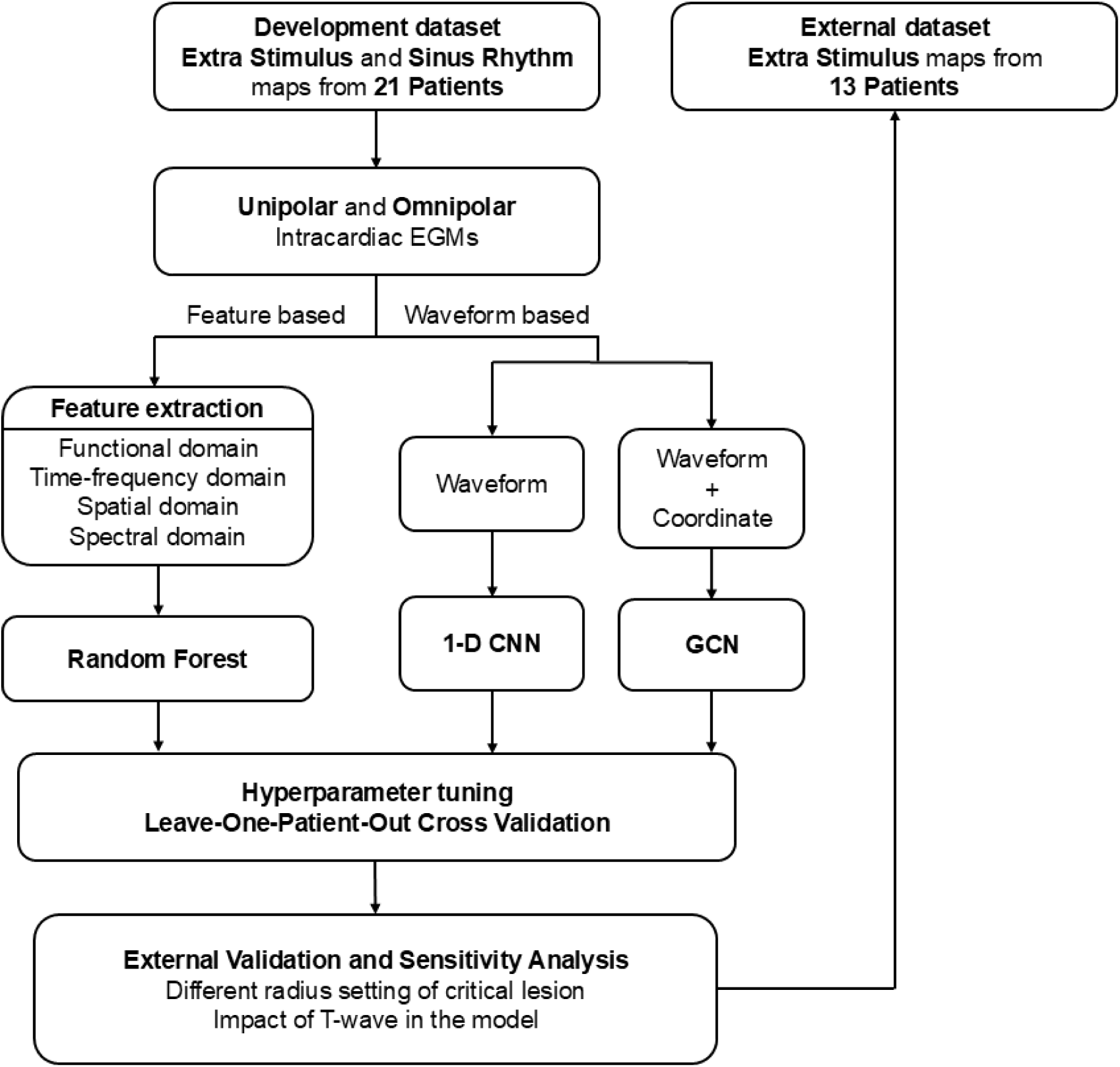
Complete workflow for this study. 1-D CNN: one-dimensional convolutional neural network, GCN: graph convolutional network.

A compact one-dimensional CNN (ResNet1D) was designed for pointwise classification of single channel intracardiac EGM waveform. The network begins with a 1-D convolution that projects the waveform to a configurable base channel dimension, followed by a sequence of same-padded 1-D convolutional blocks. Each convolutional block is followed by a rectified linear unit (ReLU) activation. Temporal information was aggregated using an adaptive average pooling layer that can reduce the time dimension to a single vector and then project to a single logit by a linear head. The class probability was calculated with a sigmoid activation. Optimisation was achieved using Adam with weight decay and a focal loss (*α* and *γ*) to mitigate class imbalance (∼10% VT ablation targets per patient) ^27^. Model hyperparameters, including channel dimensions, number of layers, kernel size, learning rate, weight decay and focal-loss parameters, were optimised with the Optuna framework.

For the GCN approach, the patient-level electroanatomical maps were represented as graphs constructed by nodes and edges. Each node corresponded to a mapping point with its node feature corresponding to the single channel EGM waveform. Undirected edges were formed by a K-nearest-neighbour graph with Euclidean distance computed from the 3D coordinates of the mapping points. The GCN model was implemented with *torch-geometric,* which consisted of multi-layer *GCNConv* blocks. This architecture projects the input node feature to a configurable hidden dimension and applies a sequence of *GCNConv* layers with ReLU after each hidden layer. The final layer produced a single logit per node, which is converted to a probability with a sigmoid activation for binary classification. The training process implemented a focal loss for class imbalance and Adam optimisation with weight decay. Hyperparameter search with Optuna, including number of neighbours K, hidden dimension, number of layers, learning rate, weight decay and focal loss parameters, was utilised.

### Evaluation and validation

Four data collection modalities, including SR-omnipolar, SR-unipolar, ES-omnipolar, and ES-unipolar, were analysed with RF, CNN and GCN, to explore the performance while using different map types (i.e. SR vs extra-stimulus pacing), signal configurations (i.e. unipolar vs omnipolar) and algorithms (i.e. RF, CNN, and GCN). Model development and evaluation followed a nested leave-one-out cross-validation (LOOCV) at patient level to emulate the clinical scenario and to obtain an unbiased estimate of performance. Validation data was z-score normalised using training-set statistics to avoid data leakage. In the LOOCV framework, across the 21 patients, each patient was sequentially held out as the outer test case. The remaining twenty patients formed the development set for model training and hyperparameter optimisation (shown in supplementary Figure 2). Within each inner LOOCV fold, one patient was designated as the inner held-out set for hyperparameter evaluation, while the remaining nineteen patients were used for model training. This nested structure can reduce the risk of optimistic bias in performance estimation by ensuring that hyperparameter tuning is performed independently of the evaluation data.

For the RF and CNN, all mapping points from a given patient were considered as dependent observations and assigned to the training or validation set according to the patient-level partitioning used in the inner LOOCV procedure. For the GCN model, each patient map was represented as a single graph, and the same patient-based LOOCV procedure was applied directly at the graph level. The training was limited to a maximum of 50 epochs, with early stopping applied using a patience of 5 epochs if the validation loss failed to decrease. Hyperparameters were tuned within the inner loop through Bayesian optimisation implemented in Optuna, with 300 trials per outer fold. After identifying the optimal hyperparameters for a given outer fold, the model was retrained using all twenty development patients and then evaluated on the held-out test patient.

For each outer held out test patient, the ability to discriminate cardiac mapping sites corresponding to VT ablation targets was assessed using the area under the ROC curve (AUC). Other metrics, including area under the precision-recall curve (average precision, AP), F1-score, precision, sensitivity and specificity, were reported based on the cut-off threshold closest to the upper-left corner of the ROC curve. Overall model performance metrics were reported using the median with interquartile range (IQR) across nested LOOCV.

For each case, localisation performance was assessed by measuring the average minimum distance between VT critical sites identified during the procedure and the predicted ablation targets (supplementary Figure 3).

### External validation

An external validation cohort, including 13 consecutive VT ablation patients with ischemic heart disease from Barts Heart Centre and St Thomas’ Hospital (London, United Kingdom), was tested ^9^. Electroanatomical maps were collected using Ensite Precision (HD Grid, Abbott, IL, USA) in 9 patients, and using CARTO (PentaRay and DecaNav, Biosense Webster Inc., CA, USA) in 4 patients, with sampling frequencies of 2034.5 Hz and 1000 Hz, respectively. Only unipolar EGMs from ES maps were available in the external dataset. In the external validation cohort, VT sites of origin were identified using standard electrophysiological techniques, including entrainment mapping for hemodynamically tolerated VTs (4 cases) and pace mapping for unstable VTs (9 cases).

All test EGMs were processed using the same windowing strategy as the development dataset for 400 ms and uniformly resampled to 2000 Hz. The median value of the hyperparameters from the LOOCV process was used to train the final ES-unipolar model using all twenty-one development patients and validated on external test patients. The patient characteristics and data processing of the external dataset were described in the supplementary Table 2.

### Sensitivity analyses

Sensitivity analyses were performed to investigate the impact of changing the size of the critical VT isthmus zone that defines the positive class (i.e. the number of mapping points considered to be VT ablation targets). The spatial definition of the critical VT isthmus zone was adjusted by changing its radius from 4 mm to 10 mm from each VT critical site. AUCs were calculated for each radius configuration through nested LOOCV.

### Data ablation analysis

In intracardiac EGMs, ventricular activation and repolarisation features are encoded in signal segments recorded during and after the QRS complex, respectively. To evaluate the importance of incorporating repolarisation features into the models, the analysis was repeated using shorter EGM segments (<250 ms) captured exclusively during ventricular activation.

Beyond differences in ventricular activation patterns, SR maps typically have a higher mapping point density than ES maps. To assess the impact of point density on model performance, analyses were repeated after down-sampling SR maps by selecting points nearest to ES mapping locations.

## Results

### Mapping characteristics

Both unipolar and omnipolar maps were collected in all 21 patients under SR and paced rhythm. The SR map showed the highest point density, in SR-omnipolar, the median of points per map was 3,982 (IQR 2,594-5,080) with a total of 80,783 points, 7,934 (9.8%) VT ablation targets were labelled (Table 2). SR-unipolar including 62,506 points with 6,601 VT ablation targets (10.6%), the median of points per map was 2,869 (2,111-3,879). The ES map showed sparser map point density, with a median of 1,368 (1,070-1,987) per map and 35,122 points in total for the ES-omnipolar map. In ES-unipolar maps, the median number of points per map was 608 (473-996) with 16,256 points in total. The proportion of VT ablation targets was higher in ES maps, with 5,082 (14.5%) and 2,296 (14.1%) for omnipolar and unipolar EGMs in total, respectively.

An example of the VT activation map and corresponding voltage map in SR is shown in **Figure *2***, along with the VT critical sites (white squares) and the VT ablation targets (< 8 mm from the critical sites).

**Figure 2.**
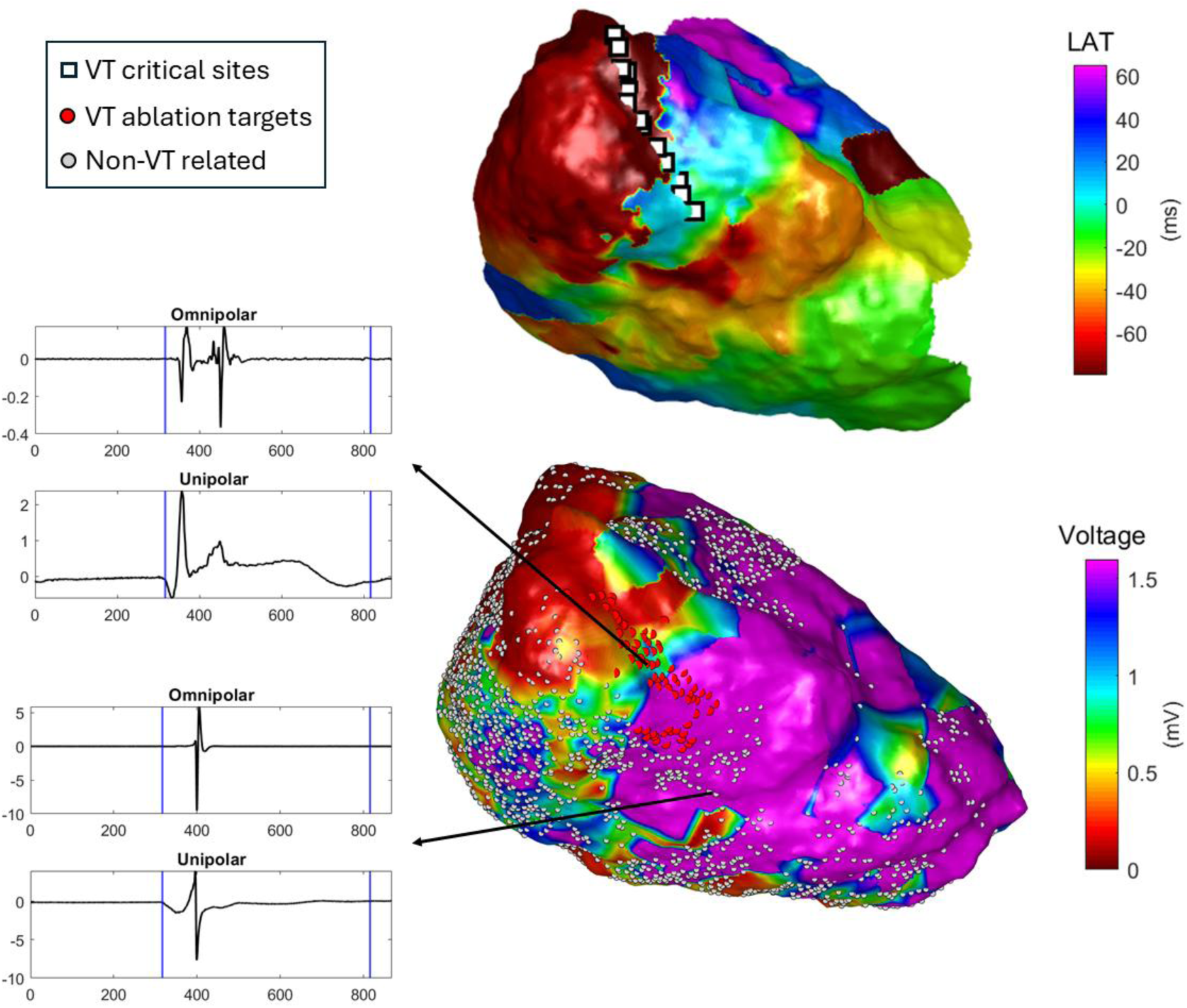
Visualisation of the VT activation map and unipolar voltage map. Top: VT local activation map, with VT critical sites marked as a white square; Bottom: unipolar voltage map from sinus rhythm, including 3,019 mapping points in total, 72 are VT ablation targets (red) with a defined radius set as 8 mm from VT critical sites, the rest of the points are marked as non-VT related (grey). Examples of omnipolar and unipolar waveforms collected from both classes are shown.

### Prediction of VT ablation targets

**Figure *3*** reports the prediction performance across three algorithms on the four data collection modalities (see Supplementary Table 3-4 for detailed patient-level results).

**Figure 3.**
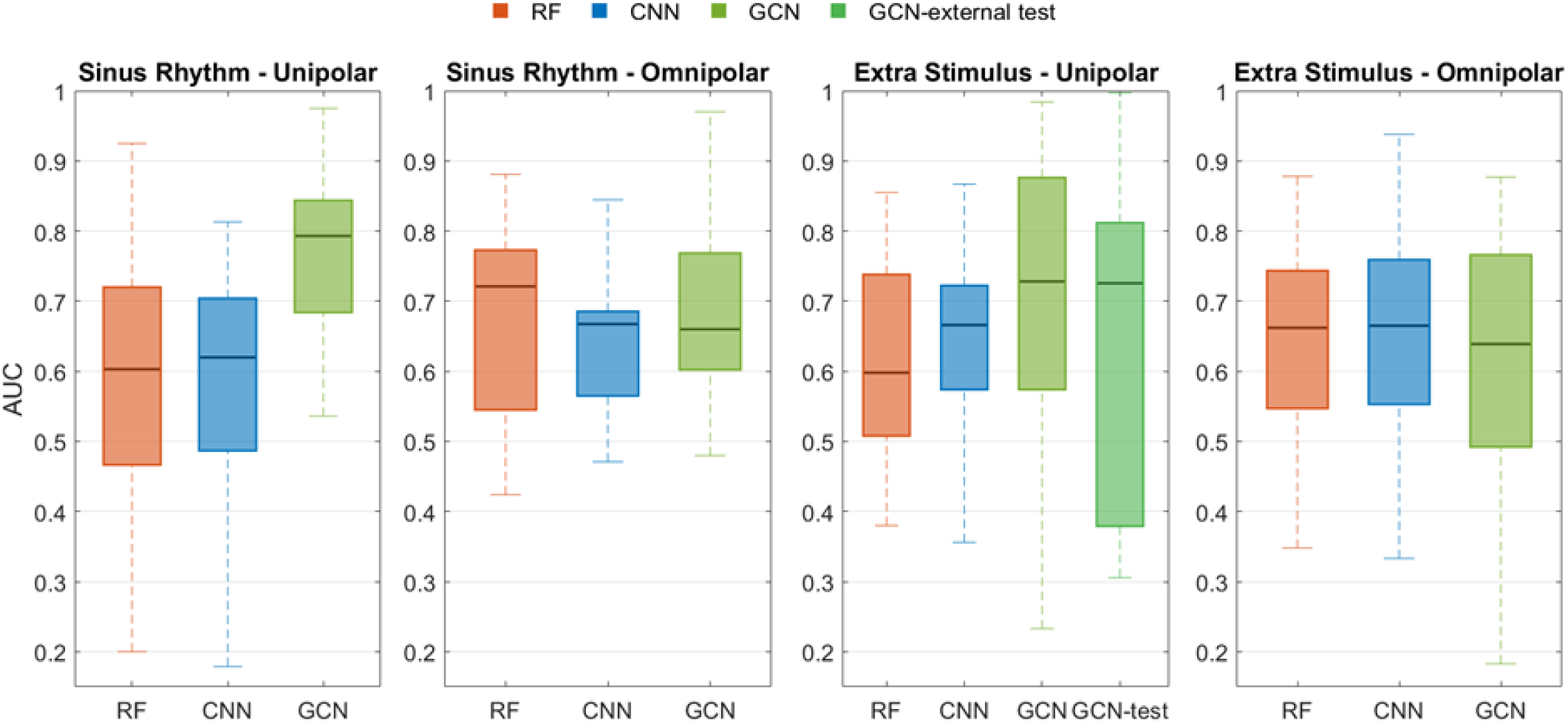
Prediction performance across algorithms and data modalities during nested LOOCV. The GCN-test is the test results on external validation dataset. Each box represents the distribution of AUCs for 21 hold-out test patients during nested LOOCV with optimized hyperparameters. The centre line indicates the median and the box edges show the IQR. Whisker extends to the observations that lie within 1.5 times the IQR below the lower quartile and above the upper quartile, values beyond this range are plotted as outliers.

Overall differences in AUC across different machine learning models were statistically significant (Friedman test, p = 0.01). The GCN achieved the highest median AUC in unipolar settings for both SR and extra stimulus models, whereas the performance difference between algorithms was smaller in the omnipolar settings and varied by data collection modality.

Among all configurations, the GCN trained on unipolar EGMs collected in SR demonstrated the highest median discrimination ability, achieving a median AUC of 0.793 (IQR: 0.68-0.84), with a sensitivity of 83.6% and specificity of 69.0%. The GCN model trained using ES-unipolar data also showed strong prediction ability, with a median AUC of 0.728 (IQR: 0.57-0.88), 82% sensitivity and 65% specificity. The detailed performance metrics for optimised GCN models are shown in Table 3.

**Table 3.**
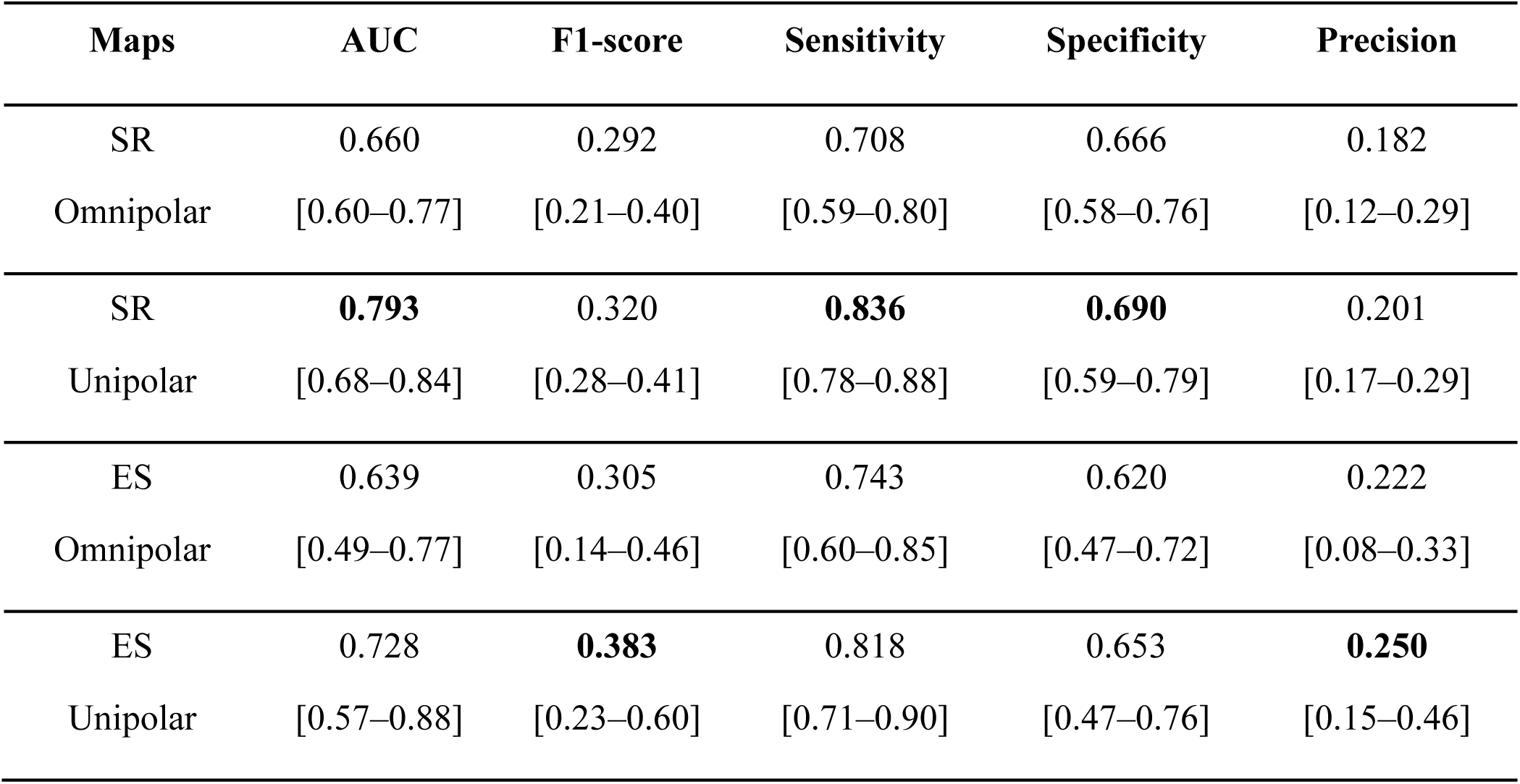
Results for GCN algorithm on different data modalities with optimal parameters across nested LOOCV. The results are reported as median [IQR].

**Figure *4*** illustrates the spatial distribution of correct and incorrect predictions on the substrate map for a representative patient (see supplementary Table 5 for detailed per patient results).

**Figure 4.**
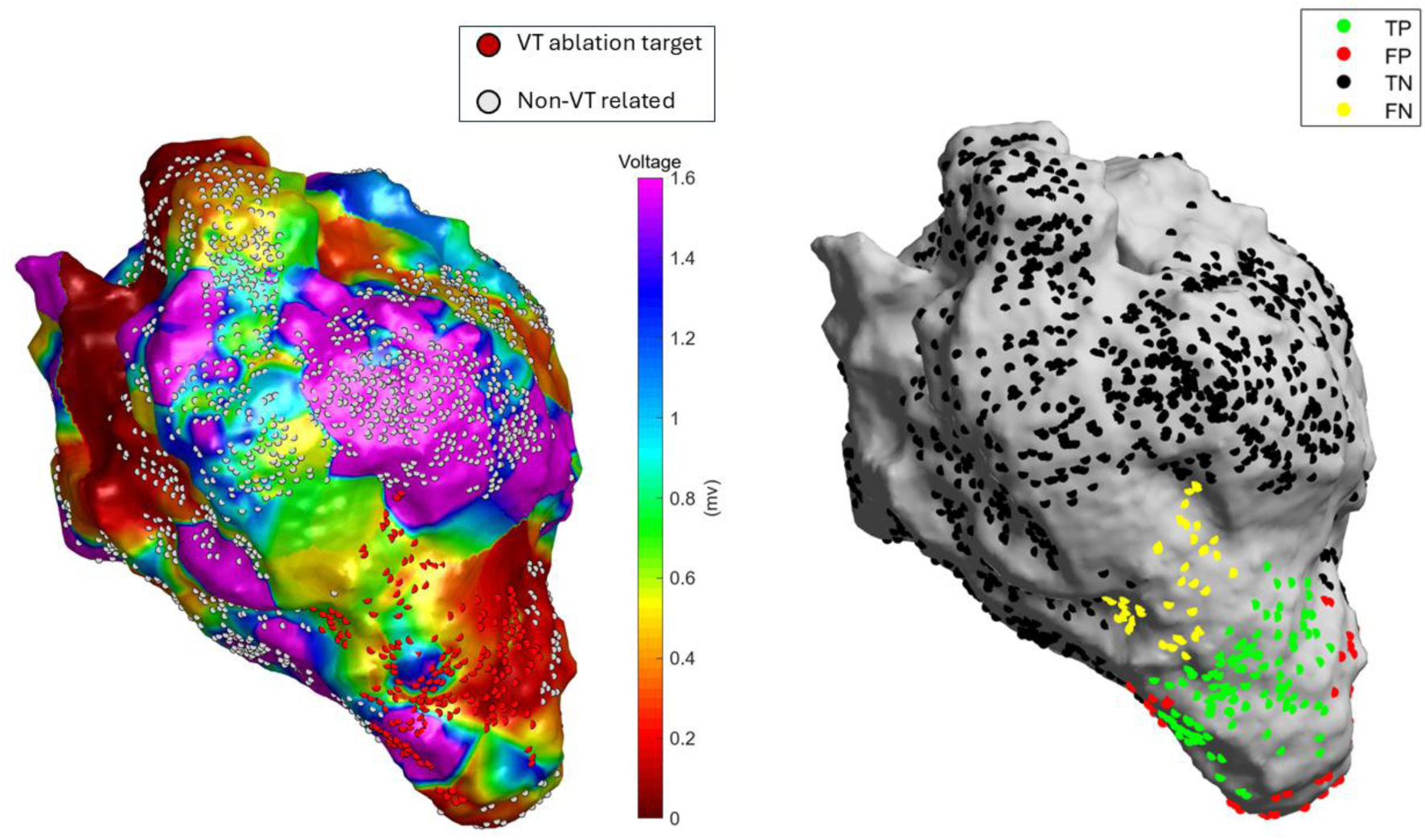
Visualisation of prediction results by GCN model using nested LOOCV. The voltage map (left) and prediction results (right) shown on 3D mesh, with class of true positive (TP), false positive (FP), true negative (TN) and false negative (FN). The AUC=0.800, F1=0.398, sensitivity=66.0%, specificity = 86.5%.

Using the GCN model on SR-unipolar data, the median distance between each VT critical site and the nearest predicted ablation target was 1.34 mm (IQR: 0.90-2.50, supplementary Table 8).

### External validation

The external validation dataset included unipolar electrograms collected during ES mapping only.

We performed external validation of the GCN model, as this was the model showing the best discrimination in the developmental model. Across 13 test patients (**Figure *3***, supplementary Table 6-7), the median test AUC was comparable to that observed in the development dataset (0.726 vs 0.728, variation of -0.3%), although the variability was slightly greater (IQR: 0.38-0.81 vs 0.57–0.88). Sensitivity and specificity also showed robust performance with a median of 88.5% and 62.4%, respectively.

### Sensitivity analyses

Varying the size of the region defining ablation targets altered class prevalence but had limited impact on model performance (**Figure *5***). For example, in SR-unipolar data, expanding the radius from 4 to 10 mm increased the median positive class proportion from 5.3% to 13.0%, while GCN performance remained stable across radius adjustments, with median AUC ranging from 0.75 to 0.80 without significant overall difference (Friedman test, p = 0.76).

**Figure 5.**
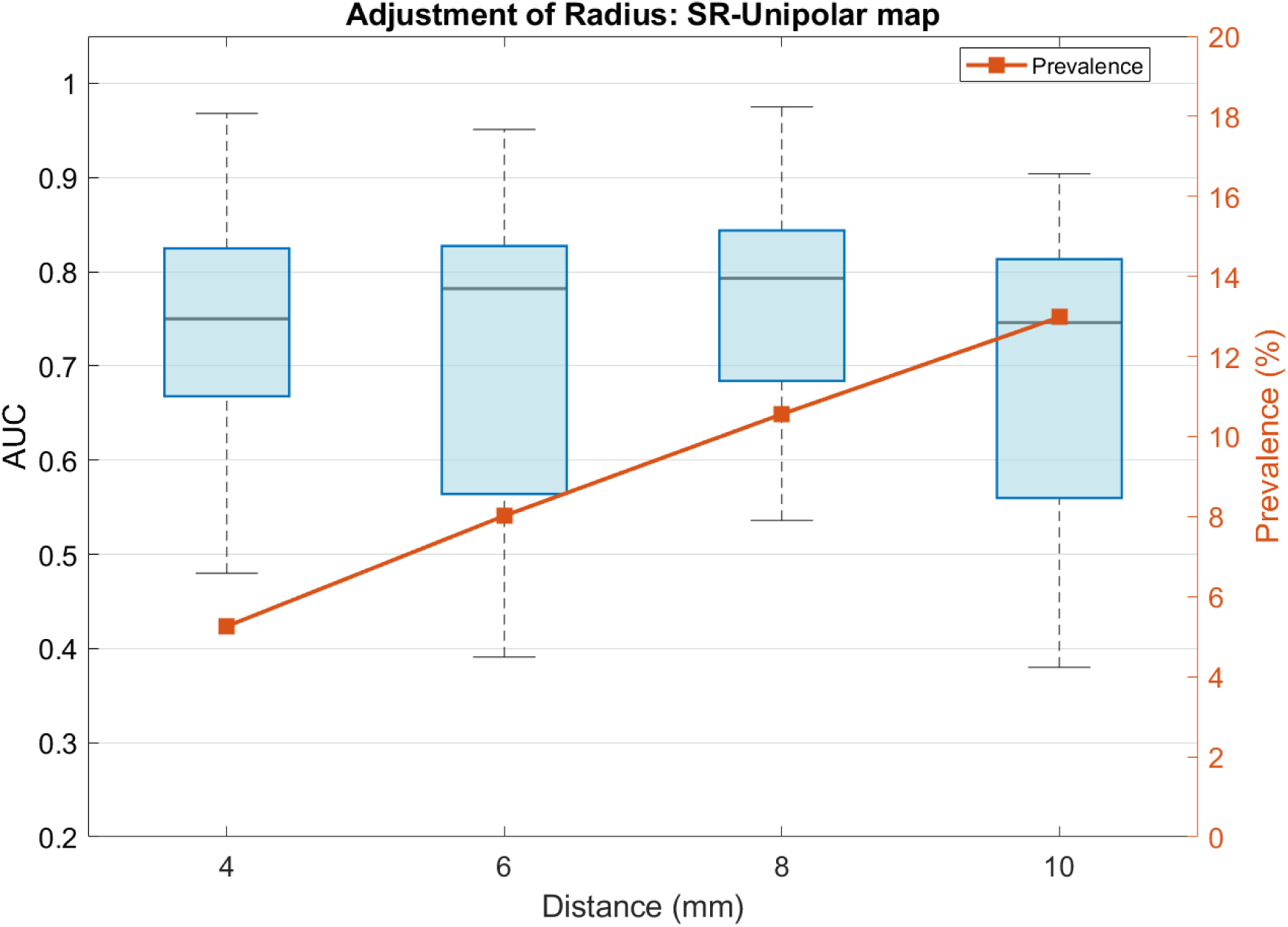
AUCs and prevalence across nested LOOCV for GCN model under different radius settings, from 4 mm to 10 mm. Boxplots show the distribution of patient-level AUCs obtained using nested LOOCV when positive labels were defined as mapping points located within 4, 6, 8, or 10 mm of the VT critical site. The orange line shows the corresponding prevalence of positive points at each radius, expressed as a percentage on the right y-axis.

### Data ablation analysis

Excluding repolarisation segments from the EGMs and training models using only ventricular activation segments had minimal impact on performance for models trained on omnipolar EGMs, but noticeably reduced performance for models trained on unipolar EGMs (Table 4). Specifically, the median AUC of GCNs trained on unipolar EGMs decreased from 0.793 to 0.718 for SR data and from 0.728 to 0.603 for ES data.

**Table 4.** Results comparison for GCN algorithm between all EGM and QRS-only segments across different data modalities by LOOCV. The results are reported as median [IQR].

| Signals | Maps | All EGM | QRS-only |
| --- | --- | --- | --- |
| Unipolar EGMs | Sinus Rhythm | 0.793<br>[0.68–0.84] | 0.718<br>[0.53–0.75] |
|  | Extra Stimulus | 0.728<br>[0.57–0.88] | 0.603<br>[0.39–0.78] |
| Omnipolar EGMs | Sinus Rhythm | 0.660<br>[0.60–0.77] | 0.712<br>[0.60–0.78] |
|  | Extra Stimulus | 0.639<br>[0.49–0.77] | 0.625<br>[0.55–0.76] |

Artificially reducing the point density of SR maps to progressively match that of ES maps, decreased GCN performance (supplementary Figure 4). The median AUC fell from 0.793 for the original SR maps to 0.729 for SR maps with twice the ES map point density, and to 0.688 for SR maps matched to ES map’s point density, which is lower than the median AUC for GCNs trained on unipolar ES electrograms (0.728).

## Discussion

The aim of this work was to validate, for the first time, the feasibility of using advanced machine learning models to automatically identify clinical VT ablation targets from EGMs collected during sinus or paced rhythm. The primary findings are: (1) Graph convolutional network outperformed CNN and feature-based models, achieving the best predictive performance on unipolar SR maps (median AUC=0.793, sensitivity=83.6% and specificity=69.0%). (2) The GCN model demonstrated robust external validity across a dataset collected using different electroanatomical mapping systems, with only -0.3% variation in median AUC. (3) Explainability analyses suggest that the superior performance of models trained on unipolar SR maps may be partly attributable to the inclusion of repolarisation information uniquely captured by unipolar EGMs and to the higher point density of SR maps, both of which improved localisation of VT critical sites.

Many novel mapping techniques and clinical metrics have been proposed and applied to guide catheter ablation for VT, but the procedure remains time-consuming and ablation outcome sub-optimal. Parallel approaches have focused on isthmuses between scar regions or corridors identified by imaging to simplify substrate ablation with the recent InEurHeart trial using CT demonstrating shorter procedure times maintaining efficacy and safety versus conventional approaches ^28,29^. However, even in this trial, if monomorphic VT was induced, a combination of activation, substrate, and/or pace-mapping was utilised highlighting the need for electrophysiological data. This study developed an automated electrogram-based solution to guide VT ablation that does not require VT induction or complex signal preprocessing. This GCN approach has the potential to assist clinicians in identifying ablation targets safely and efficiently during substrate mapping, improving ablation outcomes.

Compared to feature-based approaches such as the RF algorithm ^17^, deep learning enables end-to-end learning directly from raw EGM data, reducing reliance on manual feature engineering. Feature-based pipelines require complex pre-processing steps, including filter design, beat selection and annotation, and spectrum calculation, which are time-consuming and highly sensitive to signal quality.

Under the deep learning framework, GCN shows distinct advantages for analysing VT substrate mapping data. GCN operates on minimally processed raw EGMs exported directly from the mapping system, avoiding complex feature extraction work and reducing the risk of error propagation. By encoding each patient’s electroanatomical map as a K-nearest neighbour graph in 3D space, the GCN incorporates spatial and electrophysiological relationships between neighbouring mapping points. Specifically, the raw EGM segment recorded at each mapping point (node) is compressed into a low-dimensional feature representation, while electrophysiological information from adjacent nodes connected by edges can be aggregated through message passing. This structure allows the model to capture substrate continuity and local spatial relationships, potentially improving robustness to noise, border zone heterogeneity, and irregular sampling density. Therefore, as a node-level classification algorithm, the GCN is suited for VT ablation target prediction based on electroanatomic maps.

Despite facing challenges from limited training size and individual variability, our GCN model still demonstrated strong generalisation performance in external validation, showing only -0.3% variation in median AUC. Importantly, the external dataset was collected from a different centre, with separate clinical protocols and alternative mapping systems (CARTO) or systems with a different software version (Ensite Precision^TM^). The external dataset included only unipolar EGM collected during ES, therefore validation of our best-performing model (i.e. SR-unipolar) was not possible. Our model preserved discrimination under these heterogeneous data source conditions and provides preliminary evidence of cross-lab robustness in clinical applications. However, as differences in electrode geometry filtering, signal processing strategies, and reference template can affect model performance, further multicentre validation or centre-specific calibration are required before clinical translation.

Although substrate maps with omnipolar configuration provided higher map point density, models trained using unipolar EGMs provided better classification performance. This observation could be explained by the ability of unipolar EGMs to preserve repolarisation-related information that is attenuated or eliminated in the omnipolar configuration ^23,30^. Data ablation analysis showed a considerable reduction in classification performance when the post-QRS segments were excluded from unipolar waveforms. Late potentials and abnormal repolarisation, which both impact the morphology of the unipolar EGMs post-QRS, are considered critical indicators of slow conduction channels and arrhythmogenic substrate ^31–33^.

Programmed ES to unmask functional substrate is an increasingly utilised approach during substrate mapping. In this study, however, the GCN model trained on data collected in SR outperformed models trained on data collected during ES. These findings suggest that accurate prediction of the VT ablation target may be achievable during SR, which is faster and potentially safer than ES protocols. Explainability analyses indicated that a higher map point density in SR maps may partly account for improved performance; indeed, when the SR point density was artificially reduced to match that of ES maps, the performance superiority of SR maps was no longer observed.

Although median AUC values were relatively high, large performance variability was found between patients (supplementary Table 5). The model sensitivity to rhythm, EGM type, or lesion size can vary among patients. A possible explanation is that maps with extensive scar and multiple potential re-entry channels may benefit more from ES maps, as premature stimulation can highlight functional conduction delay and re-entry pathway, whereas maps with well-defined scar borders may be sufficiently characterised using SR maps. The number and spatial distribution of mapping points identified as ablation targets may also have an impact on the model performance. Although sensitivity analysis showed that the overall performance remained stable while increasing the number of mapping points considered as potential ablation targets, significant patient-level variability was observed. When the VT ablation target region expanded by increasing the radius from 4 to 10mm, the responses across subjects were contrasting: in one patient, the AUC increased from 0.655 to 0.799, while in another decreased from 0.822 to 0.512. This difference can be driven by patient-level heterogeneity in scar geometry and mapping quality, the radius of 6-8mm was considered as a compromise in this study.

Low F1-score and precision were observed in this study, which is an inherent problem for datasets with severe class imbalance (less than 10% of mapping points were labelled as VT ablation targets). The imbalance reflects the true clinical distribution of critical VT regions, where most tissues are non-critical to VT. In such conditions, even if our model provided high sensitivity and specificity, a large number of false positives relative to true positives predictions can lead to low F1-score and precision. However, the spatial distance analysis demonstrated that predicted VT ablation targets were often located close to true VT critical zones (median distance of 1.34 mm). This suggests that our model is able to identify regions closely approximating clinically relevant VT substrate. Developing models on a larger multicentre cohort may also improve the stability and robustness of predictive performance. Importantly, the high AUC demonstrates that the model is able to discriminate between VT ablation targets and normal sites effectively, suggesting that our model can still be a valuable tool to highlight critical VT regions.

Compared to traditional substrate-based mapping techniques, such as late potential or LAVA, the identification sensitivity for VT circuits is only about 30% ^34^, whereas GCN can provide better predictions without complex signal processing and analysis. There is limited research on methods to harness machine learning and artificial intelligence to identify VT ablation targets. Previous studies have used an ensemble tree to predict abnormal ventricular EGMs (accuracy 93.1%) and a time series convolutional neural network to identify scar on animal models (AUC > 0.90) ^15,35^. However, none of these methods can directly indicate VT ablation targets since not all abnormal potentials and scar tissue regions are related to re-entry circuits.

The GCN model achieved superior classification performance to RF and CNN by combining spatial electrophysiological properties embedded in electroanatomical maps. A multimodal algorithm could be developed by including patient demographic features and evaluation of ablation outcomes to improve patient-level robustness and prediction effectiveness. GCNs could also be used or combined with non-invasive mapping modalities such as ECG imaging and magnetic resonance imaging to offer a safe and efficient prediction of VT circuits ^36–38^.

### Limitations

The primary limitation of this study is that deep learning algorithms were developed on a relatively small cohort (21 patients). The VT mechanisms can be influenced by multiple factors, with varying infarct locations and complex scar regions among patients, it is often difficult to map the re-entry circuit in all cases. Future work should focus on establishing a larger multicentre cohort for model training and validation to improve the model’s robustness and prediction accuracy.

Although explainability analyses suggest that repolarisation contributes to prediction in the GCN model, this approach remains a ‘black box’ at the clinical level. Explainable AI approaches, such as temporal occlusion, Grad-CAM, and integrated gradients, could be applied in larger cohorts to provide mechanistic insights and link predictions to physiological mechanisms ^39–41^

## Conclusion

This study proposed the first graph-based deep learning approach to predict VT ablation targets using human electroanatomical mapping data. By analysing electrograms from substrate maps collected during SR and pacing, the model achieved good prediction accuracy for critical VT circuits in patient-level cross validation (AUC=0.793) and maintained robust performance in an independent external cohort. Further analyses provide physiological support for the deep learning algorithm, demonstrating that the GCN can automatically learn the critical substrate features supporting re-entry that are targeted by electrophysiologists during VT ablation. Indeed, it could complement or enhance imaging guided substrate ablation which is gaining momentum.

## Data Availability

All data produced in the present study are available upon reasonable request to the authors

## Acknowledgements

The authors would like to thank the clinical teams at the University Hospital Coventry, Barts Heart Centre, and St Thomas’ Hospital.

## Data availability

The data underlying this article will be shared on reasonable request to the corresponding author.

## Non-standard Abbreviations and Acronyms

ARI: Activation recovery interval
AUC: area under the receiver operating characteristic curve
CNN: Convolutional neural network
ES: Extra stimulation
GCN: Graph convolutional networks
IQR: Interquartile range
LAT: Local activation time
LOOCV: Leave-one-out cross-validation
LV: Left ventricle
ReLU: Rectified linear unit
RF: Random forest
RT: Repolarisation time
SR: Sinus rhythm
VT: Ventricular tachycardia

