## Supplemental figures and tables for "Utilising Artificial Intelligence to Identify Ventricular Tachycardia Ablation Targets in Sinus Rhythm"

**Supplemental Material**

**Model architecture and parameters**

1. **Random forest model:**

The random forest classifier was implemented using ‘RandomForestClassifier’ package in scikit-learn. The model consisted of an ensemble of decision trees trained on bootstrapped or non-bootstrapped samples, with node splitting based on randomly selected subsets of input features.

The hyperparameter search space including:

| **Parameters** | **Search space** |
| --- | --- |
| The number of trees | 100, 200, 300, 400, 500 |
| Maximum tree depth | 10, 20, 30, unrestricted |
| Minimum samples per leaf | 1, 2, 4, 8 |
| Number of features at each split | 50% of features, log2, square root |
| Bootstrap sampling | True (0.6-1.0) or false |
| Class weight | balanced or false |

2. **Convolutional neural network**

The convolutional neural network was implemented in PyTorch 2.6.0 for classification using raw electrogram waveforms as input. Each electrogram was represented as a single channel one-dimensional signal.

The model consisted of multiple sequential convolutional blocks. Each block included a one-dimensional convolution layer, batch normalisation, ReLU activation, and max pooling. The feature maps were processed through global average pooling, followed by a fully connected hidden layer, ReLU activation, a second dropout layer, and a final single neuron output layer.

The hyperparameter search space including:

| **Parameters** | **Search space** |
| --- | --- |
| Number of convolutional layers | 3-6 |
| Convolutional channels | 64-192, step size 32 |
| Convolutional kernel size | 5, 7, 9, 11 |
| Learning rate | 1e-4 to 1e-3 |
| Weight decay | 1e-6 to 1e-3 |
| Focal loss alpha | 0.5-0.85 |
| Focal loss gamma | 1-3 |

3. **Graph convolution network**

The GCN was implemented using PyTorch Geometric for node-level classification, which consisted of multiple graph convolutional layers. The first GCNConv layer projected the input electrogram feature dimension to a hidden dimension, followed by a variable number of hidden graph convolutional layers with ReLU activation. The final GCNConv layer projected the hidden representation to a single output logit per node.

The hyperparameter search space including:

| **Parameters** | **Search space** |
| --- | --- |
| Number of nearest neighbours | 10-15 |
| Hidden channels | 64-256, step size 32 |
| Number of GCN layers | 3-7 |
| Learning rate | 1e-4 to 1e-3 |
| Weight decay | 1e-6 to 1e-3 |
| Focal loss alpha | 0.5-0.85 |
| Focal loss gamma | 1-3 |

**Supplementary Figure 1**

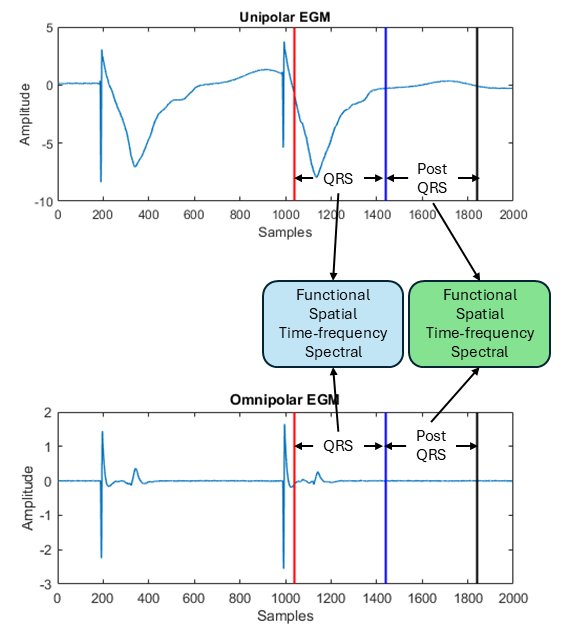

**Figure 1 Time window definition and signal segmentation.**

Red line: start point of time window. Blue line: mid-point of window. Black line: end point of time window. The window length was 400 ms in extra stimulus maps.

**Supplementary Figure 2**

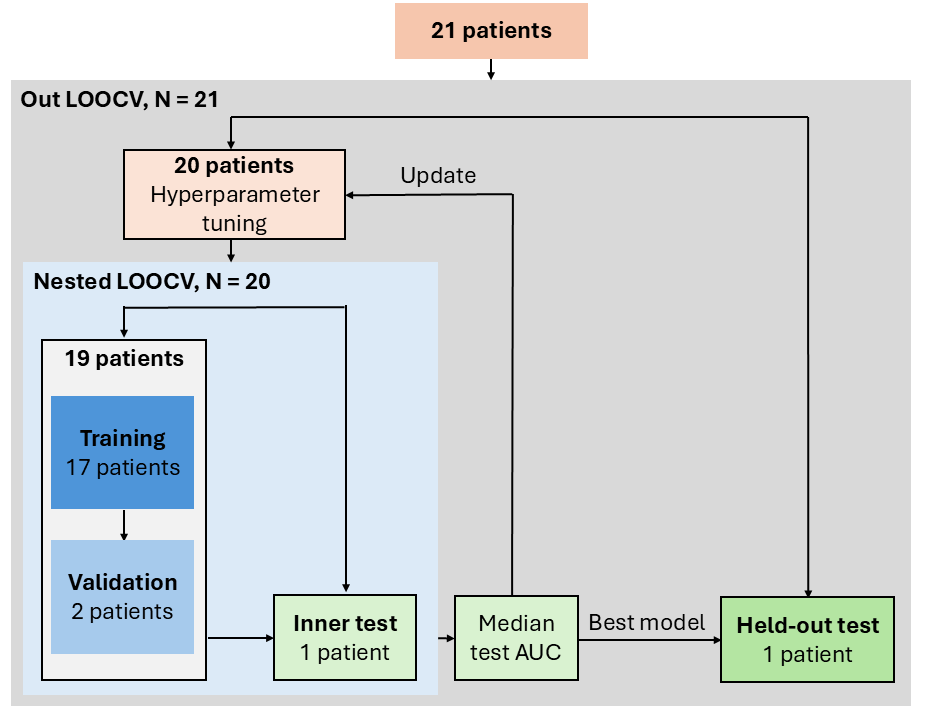

**Figure 2 Nested-LOPO cross validation procedure with hyperparameter tuning**

The grey block is the out leave one out cross validation (LOOCV) in patient level, N represents the number of iterations. In each iteration, one patient is held out as the test set, and the remaining 20 patients are used for hyperparameter tuning in the nested LOOCV (light blue block). The hyperparameters are updated only within nested LOOCV, where the held-out test patient in the outer LOOCV provides the unbiased evaluation

**Supplementary Figure 3**

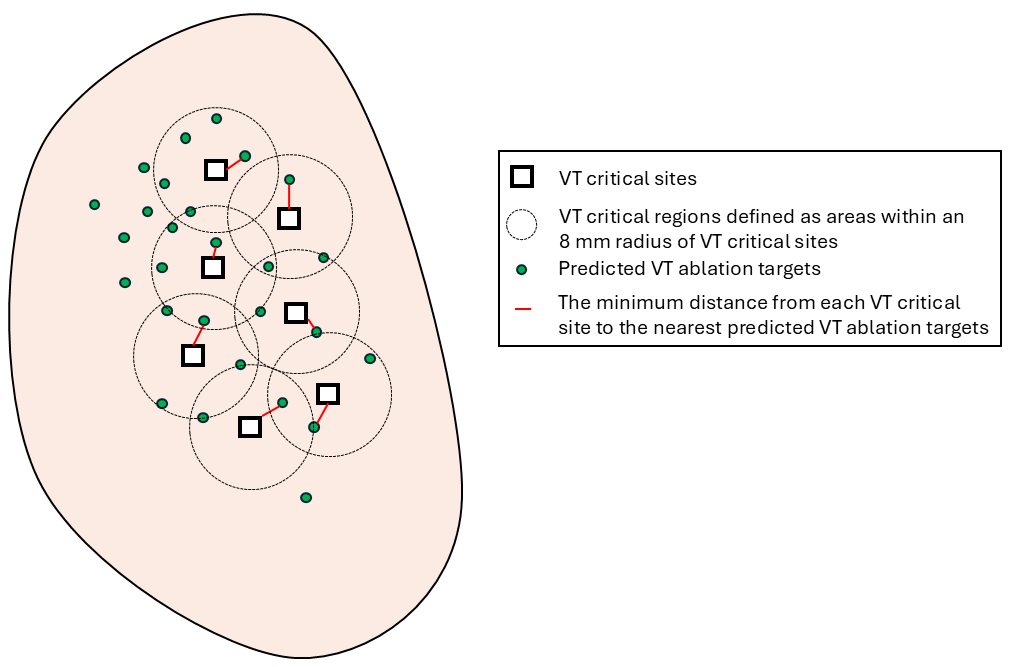

**Figure 3 The minimum distance measurement between each VT critical site and the closest predicted ablation target.**

The Black squares indicate VT critical sites identified by electrophysiologist. The grey dashed circles represent the VT isthmus zone, defined as areas within an 8 mm radius of each VT critical site. Green dots indicate the predicted VT ablation targets. Red lines show the minimum Euclidean distance from each VT critical site to its nearest predicted VT ablation target.

**Supplementary Figure 4**

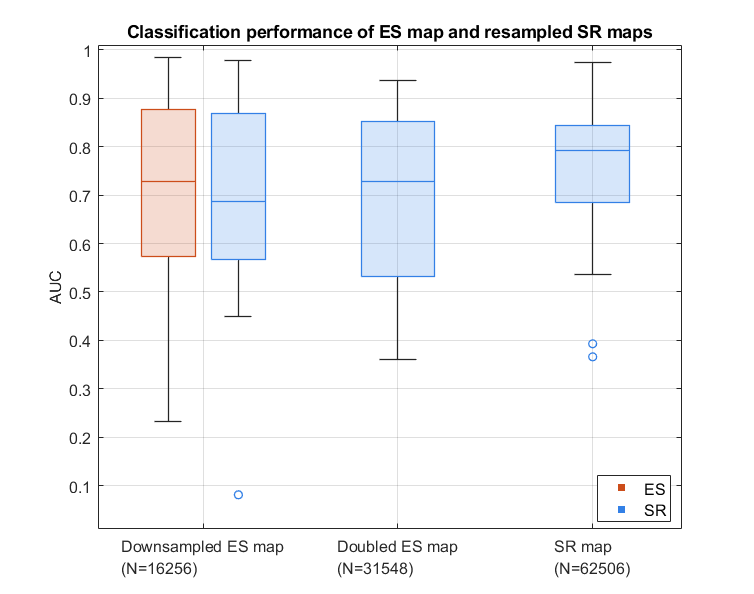

**Figure 4 Model performance of sinus rhythm unipolar model (SR-unipolar) under different mapping density conditions.**

**Down-sampled ES map**: SR-unipolar mapping points were spatially down-sampled to match the number of points in the corresponding ES map (N=16,256). **Doubled ES map**: SR-unipolar mapping points were down-sampled to match twice the ES map points (N=31,548), full SR map was used if the number of SR points was fewer than twice the ES points. **SR map**: Original SR-unipolar model without subsampling (N=62,506).

**Supplementary Table 1 Features definition.**

| **Features** | **Explanation** | **Features** | **Explanation** |
| --- | --- | --- | --- |
| ${LAT}_{O}$ | Local activation time from omnipolar EGMs | ${LAT}_{U}$ | Local activation time from unipolar EGMs |
| ${ARI}_{O}$ | Activation recovery index for omnipolar: $RT-{LAT}_{B}$ | ${ARI}_{U}$ | Activation recovery index for unipolar: $RT-{LAT}_{U}$ |
| $RT$ | Repolarization time from unipolar EGMs | $GradRT$ | Gradients of $RT$ (in ms/mm) |
| $GradAT$ | Gradients of AT (in ms/mm) | $GradARI$ | Gradients of $ARI$ (in ms/mm) |
| $P_{O}$ | Number of peaks from PSD (Over 20% of max) of omnipolar EGMs | $P_{U}$ | Number of peaks from PSD (Over 20% of max) of unipolar EGMs |
| ${Def}_{O}$ | Number of deflections within QRS of omnipolar EGMs | ${Def}_{U}$ | Number of deflections within QRS of unipolar EGMs |
| $A_{O}$ | Peak to peak voltage for Omnipolar EGMs (mV) | $A_{U}$ | Peak to peak voltage for unipolar EGMs (mV) |
| $A_{O}^{p}$ | The amplitude of omnipolar signal in post-QRS segment | $A_{U}^{T}$ | The amplitude of unipolar signal in T-wave |
| $max\vert\frac{dO_{QRS}}{dt}\vert$ | Maximum of the absolute first derivative in QRS complex of omnipolar EGMs | $max\vert\frac{dU_{QRS}}{dt}\vert$ | Maximum of the absolute first derivative in QRS complex of unipolar EGMs |
| $mean\vert\frac{dO_{QRS}}{dt}\vert$ | Mean of the absolute first derivative in QRS complex of omnipolar EGMs | $mean\vert\frac{dU_{QRS}}{dt}\vert$ | Mean of the absolute first derivative in QRS complex of unipolar EGMs |
| $max\vert\frac{dO_{p}}{dt}\vert$ | Maximum of the absolute first derivative in post-QRS for omnipolar EGMs | $max\vert\frac{dU_{T}}{dt}\vert$ | Maximum of the absolute first derivative in T-wave of unipolar EGMs |
| $mean\vert\frac{dO_{p}}{dt}\vert$ | Mean of the absolute first derivative in post-QRS for omnipolar EGMs | $mean\vert\frac{dU_{T}}{dt}\vert$ | Mean of the absolute first derivative in T-wave of unipolar EGMs |
| $D_{O}$ | Duration time for QRS in omnipolar EGMs (in ms) | $D_{U}$ | Duration time for QRS in unipolar EGMs (in ms) |
| $f_{O}$ | Central of mass frequency from the PSD for omnipolar EGMs | $f_{U}$ | Central of mass frequency from the PSD for unipolar EGMs |
| $R_{O,QRS}^{0-40}$ | Fractional energy between 0-40 Hz in QRS for omnipolar EGMs | $R_{U,QRS}^{0-20}$ | Fractional energy between 0-20 Hz in QRS for unipolar EGMs |
| $R_{O,QRS}^{40-80}$ | Fractional energy between 40-80 Hz in QRS for omnipolar EGMs | $R_{U,QRS}^{20-40}$ | Fractional energy between 20-40 Hz in QRS for unipolar EGMs |
| $R_{O,QRS}^{80-120}$ | Fractional energy between 80-120 Hz in QRS for omnipolar EGMs | $R_{U,QRS}^{40-60}$ | Fractional energy between 40-60 Hz in QRS for unipolar EGMs |
| $R_{O,QRS}^{120-160}$ | Fractional energy between 120-160 Hz in QRS for omnipolar EGMs | $R_{U,QRS}^{60-80}$ | Fractional energy between 60-80 Hz in QRS for unipolar EGMs |
| $E_{O,QRS}^{0-160}$ | Total energy between 0-160 Hz in QRS for omnipolar EGMs | $E_{U,QRS}^{0-80}$ | Total energy between 0-80 Hz in QRS for unipolar EGMs |
| $R_{O,P}^{0-40}$ | Fractional energy between 0-40 Hz in post-QRS for omnipolar EGMs | $R_{U,T}^{0-20}$ | Fractional energy between 0-20 Hz in T-wave for unipolar EGMs |
| $R_{O,P}^{40-80}$ | Fractional energy between 40-80 Hz in post-QRS for omnipolar EGMs | $R_{U,T}^{20-40}$ | Fractional energy between 20-40 Hz in T-wave for unipolar EGMs |
| $R_{O,P}^{80-120}$ | Fractional energy between 80-120 Hz in post-QRS for omnipolar EGMs | $R_{U,T}^{40-60}$ | Fractional energy between 40-60 Hz in T-wave for unipolar EGMs |
| $R_{O,P}^{120-160}$ | Fractional energy between 120-160 Hz in post-QRS for bipolar EGMs | $R_{U,T}^{60-80}$ | Fractional energy between 60-80 Hz in T-wave for unipolar EGMs |
| $E_{O,P}^{0-160}$ | Total energy between 0-160 Hz in post-QRS for omnipolar EGMs | $E_{U,T}^{0-80}$ | Total energy between 0-80 Hz in T-wave for unipolar EGMs |

**Supplementary Table 2 Distribution of mapping points in external validation patient cohort.**

Thirteen patients undergoing catheter ablation for ventricular tachycardia (VT) due to ischaemic heart disease were prospectively enrolled. All participants provided informed consent and received local ethical approval (Barts Heart Centre and St Thomas' Hospital). Only patients whose VT origin could be localized and with sufficient density in substrate map were included in the analysis.

Hemodynamically tolerable VT was confirmed by entrainment or ablation termination, while intolerant VT was confirmed by pace-mapping, requiring a ≥90% correlation between the 12-lead ECG and VT morphology. Pacing was performed at the right ventricular apex, preferably with an S1 train coupled with a short S2 stimulus to induce slowed conduction. Single pacing after the sinus R wave was used when necessary.

|  | S1S2 map | ES map | Total |
| --- | --- | --- | --- |
| Number of maps | 4 | 9 | 13 |
| Sampling frequency | 1000Hz | 2034.5Hz |  |
| Mapping points per map | 2,072  (934-3,136) | 667  (447-1,270) | 719  (447-1,841) |
| Total mapping points | 8,128 | 10,038 | 18,176 |
| VT ablation targets per map (R=8) | 29  (10-99) | 23  (15-47) | 23  (14-47) |
| Percentage of VT ablation targets per map (R=8) | 3.63%  (0.6%-6.4%) | 3.42%  (1.6%-7.2%) | 3.42%  (1.2%-6.7%) |
| Total VT ablation targets | 216 | 519 | 735 |

**Supplementary Table 3 Performance of RF algorithm with hyperparameter during nested leave-one-patient-out cross validation.**

| **Maps** | **AUC** | **AP** | **F1-score** | **Sensitivity** | **Specificity** | **Precision** |
| --- | --- | --- | --- | --- | --- | --- |
| SR  Omnipolar | 0.721  [0.55–0.77] | 0.162  [0.08–0.30] | 22.7  [14.3–38.6] | 70.8  [58.2–75.3] | 61.2  [55.1–68.9] | 13.8  [7.9–27.2] |
| SR  Unipolar | 0.603  [0.47–0.72] | 0.104  [0.07–0.21] | 22.3  [15.5–32.0] | 75.1  [64.2–80.8] | 51.7  [45.0–63.7] | 13.0  [8.6–21.5] |
| ES  Omnipolar | 0.662  [0.55–0.74] | 0.187  [0.11–0.30] | 31.6  [21.0–43.5] | 70.6  [62.2–81.1] | 61.2  [47.8–68.7] | 21.2  [12.2–31.2] |
| ES  Unipolar | 0.598  [0.51–0.74] | 0.194  [0.14–0.35] | 29.4  [19.1–44.3] | 65.2  [58.5–79.8] | 60.1  [46.8–68.1] | 19.1  [12.4–34.2] |

**Supplementary Table 4 Performance of CNN algorithm with hyperparameter during nested leave-one-patient-out cross validation.**

| **Maps** | **AUC** | **AP** | **F1-score** | **Sensitivity** | **Specificity** | **Precision** |
| --- | --- | --- | --- | --- | --- | --- |
| SR  Omnipolar | 0.667  [0.57–0.69] | 0.127  [0.08–0.26] | 19.1  [12.8–32.7] | 75.1  [63.1–84.1] | 54.5  [40.4–68.4] | 12.0  [7.0-23.1] |
| SR  Unipolar | 0.591  [0.48–0.71] | 0.118  [0.06–0.19] | 22.6  [12.8–30.8] | 71.9  [61.4–79.9] | 58.3  [41.9–60.2] | 13.6  [7.1-20.3] |
| ES  Omnipolar | 0.665  [0.55–0.76] | 0.229  [0.10–0.43] | 31.7  [16.8–47.3] | 71.4  [61.8–75.6] | 59.0  [51.4–69.7] | 21.7  [9.6-36.0] |
| ES  Unipolar | 0.666  [0.57–0.72] | 0.228  [0.15–0.37] | 37.2  [27.7–43.1] | 74.2  [63.9–78.5] | 62.6  [50.6–69.1] | 25.0  [17.4-32.5] |

**Supplementary Table 5 The GCN prediction performance per patient based on SR unipolar EGMs in the development dataset during nested leave-one-patient-out cross validation.**

| **Patient** | **Prevalence**  (%) | **AUC** | **AP** | **F1-score**  (%) | **Precision**  (%) | **Sensitivity**  (%) | **Specificity**  (%) |
| --- | --- | --- | --- | --- | --- | --- | --- |
| P1 | 4.23 | 0.88 | 0.146 | 38.1 | 23.6 | 100.0 | 85.7 |
| P2 | 7.07 | 0.828 | 0.285 | 26.2 | 15.6 | 82.7 | 65.9 |
| P3 | 8.73 | 0.717 | 0.131 | 28.7 | 17.2 | 87.7 | 59.6 |
| P4 | 14.21 | 0.874 | 0.567 | 51.4 | 37.1 | 83.6 | 76.5 |
| P5 | 2.63 | 0.843 | 0.076 | 16.9 | 9.4 | 87.5 | 77.2 |
| P6 | 4.56 | 0.536 | 0.06 | 11.1 | 6.3 | 50.6 | 63.7 |
| P7 | 8.54 | 0.788 | 0.167 | 31.7 | 19.7 | 81.2 | 69.0 |
| P8 | 7.52 | 0.800 | 0.603 | 39.8 | 28.4 | 66.0 | 86.5 |
| P9 | 10.12 | 0.802 | 0.215 | 38.3 | 24.8 | 83.3 | 71.6 |
| P10 | 26.11 | 0.366 | 0.198 | 32.1 | 24.3 | 47.5 | 47.7 |
| P11 | 8.15 | 0.786 | 0.158 | 34.6 | 21.6 | 87.2 | 71.9 |
| P12 | 25.66 | 0.975 | 0.939 | 87.1 | 80.3 | 95.2 | 92.0 |
| P13 | 6.45 | 0.701 | 0.095 | 21.7 | 12.4 | 86.6 | 57.8 |
| P14 | 1.86 | 0.95 | 0.146 | 32.0 | 19.1 | 98.9 | 92.0 |
| P15 | 10.03 | 0.393 | 0.076 | 18.6 | 10.5 | 79.9 | 24.3 |
| P16 | 14.76 | 0.633 | 0.178 | 31.5 | 20.1 | 73.1 | 49.7 |
| P17 | 14.13 | 0.593 | 0.155 | 30.6 | 19.7 | 68.0 | 54.5 |
| P18 | 19.87 | 0.793 | 0.343 | 55.5 | 40.6 | 87.6 | 68.2 |
| P19 | 11.81 | 0.831 | 0.261 | 49.0 | 34.5 | 84.4 | 78.5 |
| P20 | 16.20 | 0.767 | 0.275 | 45.1 | 30.9 | 83.1 | 64.1 |
| P21 | 4.64 | 0.846 | 0.125 | 30.9 | 18.3 | 99.0 | 78.5 |

**Supplementary Table 6 Test results on 13 external patients using pre-trained GCN model on the development dataset (extra stimulation map, unipolar EGMs).**

| **Metrices** | **Median (IQR)** |
| --- | --- |
| AUC | 0.726 (0.379–0.812) |
| F1-score | 0.148 (0.037–0.452) |
| Precision | 0.084 (0.019– 0.303) |
| Sensitivity | 0.885 (0.785– 0.917) |
| Specificity | 0.623 (0.354– 0.770) |

**Supplementary Table 7 Patient-level test results on external validation dataset using pre-trained GCN model on the development dataset (extra stimulation map, unipolar EGMs).**

| **Patient** | **System** | **AUC** | **F1** | **Precision**  **(%)** | **Sensitivity**  **(%)** | **Specificity**  **(%)** |
| --- | --- | --- | --- | --- | --- | --- |
| External 1 | CARTO | 0.997 | 0.703 | 54.9 | 97.5 | 99.1 |
| External 2 | CARTO | 0.726 | 0.234 | 13.5 | 88.5 | 62.4 |
| External 3 | CARTO | 0.927 | 0.448 | 30.0 | 88.2 | 85.4 |
| External 4 | CARTO | 0.395 | 0.002 | 0.1 | 50.0 | 37.8 |
| External 5 | Precision | 0.333 | 0.082 | 4.3 | 88.9 | 30.1 |
| External 6 | Precision | 0.755 | 0.164 | 9.0 | 90.0 | 72.0 |
| External 7 | Precision | 0.405 | 0.039 | 2.0 | 73.9 | 36.7 |
| External 8 | Precision | 0.754 | 0.025 | 1.3 | 80.0 | 74.6 |
| External 9 | Precision | 0.306 | 0.131 | 7.0 | 96.2 | 24.5 |
| External 10 | Precision | 0.933 | 0.480 | 32.3 | 93.3 | 84.1 |
| External 11 | Precision | 0.330 | 0.029 | 1.5 | 80.0 | 31.5 |
| External 12 | Precision | 0.480 | 0.148 | 8.4 | 64.7 | 46.0 |
| External 13 | Precision | 0.774 | 0.466 | 31.3 | 91.1 | 71.1 |

**Supplementary Table 8 Patient-level mean minimum distance from VT critical sites to the nearest predicted ablation targets.**

Mean ± standard deviation (SD) of the minimum Euclidean distance from each VT critical site to the nearest predicted ablation target (including both true positive and false positive points). Distance is expressed in millimetres.

| **Patient** | **Number of VT critical sites** | **Mean ± SD** |
| --- | --- | --- |
| P1 | 6 | 5.43 ± 1.84 |
| P2 | 10 | 2.11 ± 0.87 |
| P3 | 6 | 2.14 ± 2.04 |
| P4 | 14 | 0.91 ± 0.55 |
| P5 | 13 | 2.44 ± 2.58 |
| P6 | 5 | 7.31 ± 6.05 |
| P7 | 5 | 0.95 ± 0.54 |
| P8 | 15 | 4.54 ± 3.91 |
| P9 | 65 | 0.92 ± 0.70 |
| P10 | 52 | 3.31 ± 3.68 |
| P11 | 8 | 0.85 ± 0.45 |
| P12 | 46 | 2.23 ± 1.76 |
| P13 | 1 | 0.36 ± 0.00 |
| P14 | 1 | 1.05 ± 0.00 |
| P15 | 119 | 1.34 ± 0.88 |
| P16 | 33 | 1.64 ± 1.47 |
| P17 | 16 | 2.70 ± 2.93 |
| P18 | 20 | 0.85 ± 0.47 |
| P19 | 9 | 1.02 ± 1.03 |
| P20 | 27 | 0.63 ± 0.72 |
| P21 | 2 | 0.49 ± 0.21 |
